# AIM-PrEP: AI Agent-Driven Multicenter Intervention to Improve PrEP Adherence and Health Monitoring Among Men Who Have Sex with Men (MSM) – Protocol of A Randomized Controlled Trial

**DOI:** 10.64898/2026.06.02.26354777

**Authors:** Rongbiao Zeng, Zonglin Zuo, Haihang Yu, Yujing Jin, Yingxin Wang, Hui Lv, Guoyong Wang, Na Zhang, Haifeng He, Xiaojie Huang, Xingliang Zhang, Qiru Su, Junjie Xu

## Abstract

**Background:** Pre-exposure prophylaxis (PrEP) has demonstrated a significant reduction in HIV infections among men who have sex with men (MSM), however, low medication adherence hinders its preventative effectiveness. Traditional approaches, such ashealth education and face-to-face inquiry (HEF), have demonstrated certain efficacy in improving PrEP adherence. However, these methods are resource-intensive and often plagued by delays, rendering timely and precise interventions challenging. This randomized controlled trial aims to assess the effectiveness of an intervention comprising AI-chatbot for PrEP (PrEP-bot) and Smart pillbox (SPB) (PrEP-bot-SPB) strategy to improve PrEP adherence among MSM compared to HEF.

**Methods and analysis:** A three-arm, multicenter, open-lable RCT will be conducted with Chinese MSM ≥18 years. A total of 300 participants will be recruited through three sources, including hospitals, community-based organizations (CBOs) and peer referral in five cities: Shenzhen, Beijing, Qingdao, Hangzhou and Zhengzhou. After completing baseline survey, participants will be randomized evenly into interventions or control groups: the PrEP-bot group, the PrEP-bot-SPB group, and the HEF control group. Participants in the PrEP-bot group will be granted access to an AI-chatbot agent through WeChat. This agent will: 1) generate personalized PrEP medication plans; 2) provide medication reminders and PrEP-related health check-ups notifications; 3) inquire about missed doses to deliver tailored interventions; 4) answer participant questions about PrEP using guideline-based knowledge. Participants in the PrEP-bot-SPB group will receive both the SPB and the PrEP-bot interventions. SPB could delivers medication reminders. Participants in HEF group will receive a health education pamphlet introducing PrEP and knowledge related to PrEP medication adherence at baseline and face-to-face inquiry every three months. Outcomes will be assessed for both short-term and medium-to-long-term effects. The primary objective is the effectiveness in improving PrEP adherence measured by self-report, Eight-Item Morisky medication adherence scale (MMAS-8) and concentration of Tenofovir in dried blood spots (DBS) (PrEP adherence ≥90%) at 3 months follow-up. Secondary outcomes include: 1) effectiveness in preventing HIV infection measured by HIV-self test (HIVST); 2) effectiveness of PrEP-related health check-ups; 3) the effectiveness, feasibility, acceptability, and user satisfaction with the PrEP-bot; 4) effectiveness in improving PrEP adherence at 6-month, 9-month and 12-month follow-up periods. All participants will receive quarterly follow-up visits during the 12-month study period. Intention-to-treat analysis and per protocol set (PPS) analysis will be used.

**Results:** Recruitment and enrollment of participants began in January 2026 and is currently ongoing.

**Discussion:** This study is expected to establish a novel AI-based intervention model for PrEP, providing innovative strategies for HIV control among MSM populations. If the PrEP-bot is proven non inferiority than HEF, it could offer users real-time, precise, and personalized interventions while simultaneously addressing PrEP-related inquiries and health check-ups reminders. Importantly, this approach would achieve significant reductions in resource requirements for implementation and maintenance and being more cost-effective. With the ongoing advancement of AI technologies, PrEP-bot hold substantial promise for widespread implementation in PrEP adherence, potentially revolutionizing HIV prevention for MSM in China through this innovative intervention modality.

**Trial registration:** *ChiCTR2500111280* (Chinese Clinical Trial Registry). Date of registration: 29 October 2025.

## Introdction

### Background

Human immunodeficiency virus (HIV) infection remains a significant public health. According to the latest data released by UNAIDS, approximately 40.8 million people were living with HIV in 2024, with 1.3 million new HIV infections occurring that year(1). Men who have sex with men (MSM) are a key population for HIV prevention both domestically and internationally. By the end of 2024, there were a total number of 1.355 million people infected with HIV/AIDS in China, with 101,000 new cases reported. Among them, the proportion of MSM sexual transmission rose from 9.1% in 2009 to 25.2% in 2024(2). This indicates that the prevention and control measures taken for MSM in the past 40 years need to be improved.

Pre-exposure prophylaxis (PrEP) is recognized as highly effective in preventing new HIV infections in high-risk populations(3–7). Using PrEP in high-risk populations can effectively curb the spread of HIV at the public health level(6, 8, 9). The World Health Organization (WHO) guidelines also recommend that it should promote PrEP use among key populations with new HIV infection rates exceeding 3 per 100 person-years to prevent HIV infection(10, 11). By the end of 2023, 152 countries and territories have taken this suggestion(12).

However, the effectiveness of PrEP in preventing HIV infection depends on the adherence to PrEP (13). A systematic review and meta-analysis found that HIV prevention effect can reach 98.8% if PrEP adherence is good (3). However, no significant HIV prevention effect can be observed when the PrEP adherence below 40% (14). Therefore, adherence intervention must be strengthened to ensure the HIV prevention effect.

Traditional intervention measures for PrEP adherence, like face to face health education (HE), face-to-face inquiry, mainly consist of post-hoc reminders, Healthcare professionals usually assess PrEP adherence through face-to-face inquiries during follow-up. Post-hoc interventions are implemented only after PrEP participants have missed doses for weeks or months. Although these interventions can effectively improve PrEP adherence compare to no intervention, they are characterized by delayed intervention timing, fail to provide real-time support, and are both labor-intensive and economically costly(15). There is an urgent need to develop a new type of PrEP adherence intervention that is economical, convenient and can remotely implement real-time PrEP adherence intervention for PrEP users.

With the development of artificial intelligence (AI) and Internet technology, a number of clinical studies have begun to adopt new remote digital intervention measures, including mobile phone software, phone and SMS reminders, online interviews, mobile games, wechat video, etc., and have achieved initial results(16–20). Although telephone and SMS reminders can prompt individuals who have forgotten to take their medication, the messages lack personalization and have limited impact on those with low medication adherence motivation. Moreover, frequent reminder messages may lead to fatigue and aversion among individuals who already adhere well to their medication schedule, and there are potential risks of personal privacy breaches. In recent years, such methods have been seldom used by researchers for PrEP adherence interventions(21). Smart pill box (SPB), a real-in-time monitoring equipment, offer real-time adherence monitoring. It was proven to have higher effectiveness in improving PrEP adherence than traditional intervention(22). However, these measures fail to provide precise and targeted interventions to enhance PrEP adherence for MSM. Moreover, frequent reminders for those with good medication adherence can also cause fatigue and aversion. A more precise and personalized approach is needed to intervene in PrEP adherence among MSM.

The increasing prominence of large language models (LLM) has fueled growing interest in AI-chatbots which based on generative artificial intelligence (Gen-AI). AI-chatbots simulate human interaction through modalities such as text, language, and visual communication, enabling user engagement in conversational exchanges(16). Emerging research underscores the potential of AI-chatbot-assisted interventions as a valuable tool in healthcare(23–25). Two recent RCT studies have demonstrated that PrEP-bot can increase influenza vaccination rates among older adults through personalized information tailored by PrEP-bots(26) [29], as well as effectively address mental health issues in adults(27). The AI-chatbot is potentially useful for PrEP adherence for MSM. To our knowledge, this approach is innovative and has not been previously reported.

While consistent PrEP use is highly effective in preventing HIV infection, it does not protect against other sexually transmitted infections (STIs) such as syphilis, gonorrhea, and chlamydia. It can also potentially impact liver and kidney function. Therefore, regular STI screening and liver/kidney function monitoring are particularly important for PrEP users(27). However, previous studies have found that health check-up rates among PrEP users are low, falling far short of the recommended testing frequency (28, 29). So far, there is still a lack of effect evaluation on the intervention of health check-up behaviors of HIV PrEP participants.

This study aims to evaluate whether a precision intervention combining an AI chatbot for PrEP adherence (PrEP-bot) with a SPB (PrEP-bot-SPB) differs from traditional PrEP interventions [HE and face-to-face inquiry (HEF)] in improving PrEP adherence and PrEP-related health check-up behaviors among MSM. The research hypothesis is that the PrEP-bot-SPB will be non-inferior to the traditional intervention in enhancing PrEP adherence and promoting related health check-ups, while also being more cost-effective.

### Objectives

The primary objective of this study is to evaluate the difference in PrEP adherence improvement between the PrEP-bot-SPB intervention group and the HEF intervention group at 3th follow up.

The secondary objectives include:

1) exploring whether the PrEP-bot groups produce effects comparable to HEF intervention in improving PrEP adherence, respectively;
2) comparing the differences in effectiveness for preventing HIV infection and PrEP-related health check-ups rates among three groups;
3) evaluating the effectiveness, feasibility, acceptability, and user satisfaction of the PrEP-bot;
4) assessing the consistency between the proportion of good PrEP adherence (adherence 90%) based on self-report and the 8-item Morisky Medication Adherence Scale (MMAS-8) with the proportion of good PrEP adherence (TFV ≥35.5 ng/mL) based on dried blood spot (DBS) measurement and venous blood;
5) Exploring the interventions effects of the PrEP-bot-SPB and the PrEP-bot on daily PrEP and event-driven PrEP, respectively.
6) Evaluate the health economics effect of the PrEP-bot-SPB intervention compared with the traditional PrEP intervention approach.

## Methods

### Study Design

A three arms, multicenter, open-lable RCT will be conducted in January 2026. A total of 300 MSM will be enrolled on-site and randomly divided into three groups in a 1:1:1 ratio.Intervention groups are: 1) the PrEP-bot group receiving only a PrEP-bot intervention, 2) the PrEP-bot-SPB group receiving both SPB and PrEP-bot interventions. The control group receiving a health education pamphlet introducing PrEP and knowledge related to PrEP medication adherence at baseline and face-to-face inquiries. Participants will be prospectively followed for 12 months, with once clinic visits every 3 months. The outcomes will be assessed for short-term effects and short-term, medium-to-long-term effects. The SPIRIT recommendations detail is shown in supplementary file.

### Participant and Recruitment

Inclusion criteria are: (1) assigned male at birth; (2) reported anal sex with a man; (3) aged 18–65 years; (4) currently using PrEP; (5) self-reported HIV-negative status confirmed by a fourth-generation HIV test; (6) fully understand the study’s purpose, procedures, and methods, voluntarily provide written informed consent, and are willing to participate in regular follow-up visits.

Exclusion criteria: 1) Those who are unable to use the electronic pillbox at home; 2) Those with mental or psychological disorders; 3) Those unable to operate a smartphone; 4) Other situations deemed unsuitable for participation by the researchers. A flowchart diagram is shown in figure 1.

**Figure 1.**
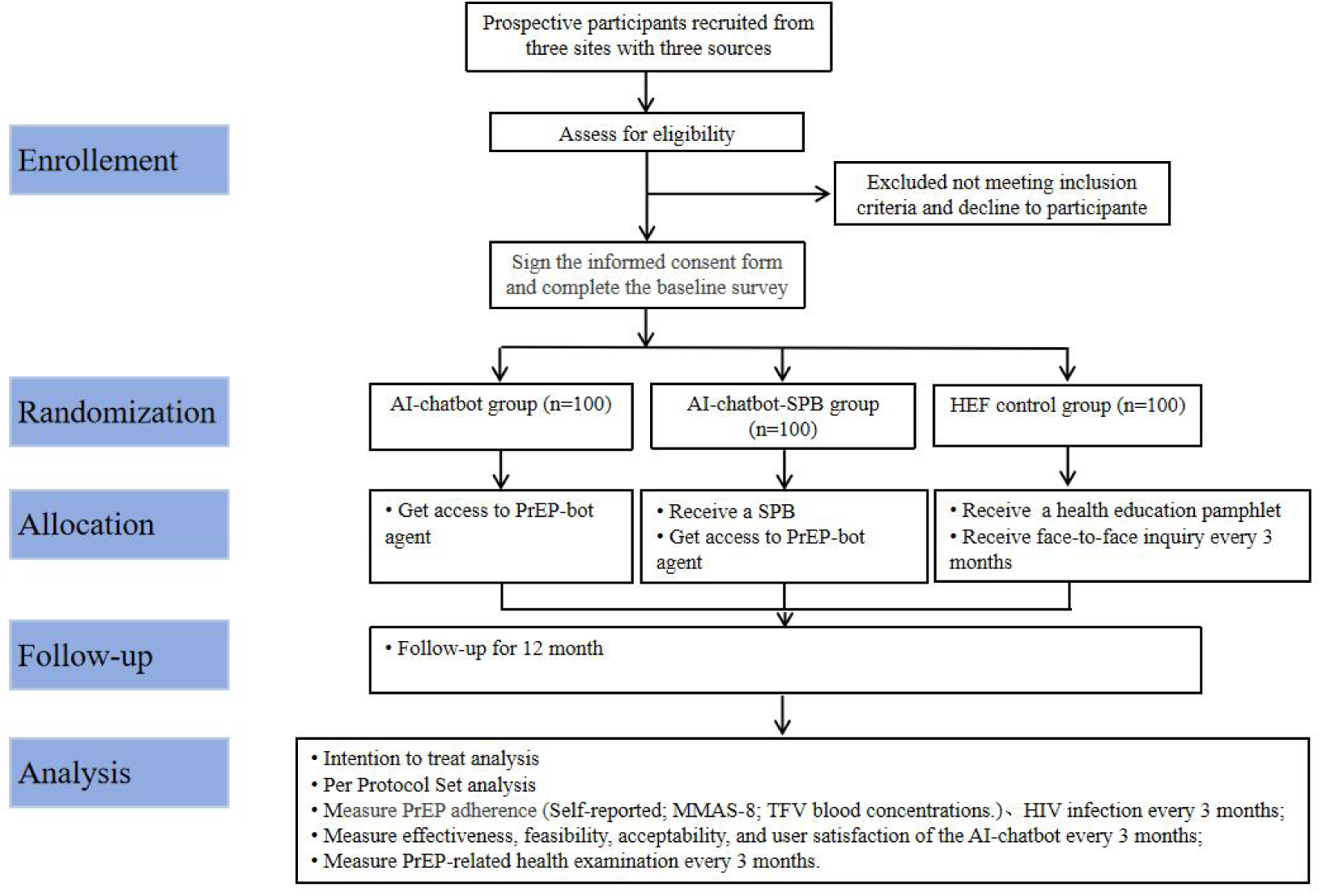
Flowchart diagram for the noninferiority randomized controlled trail. SPB: Smart pillbox; HEF: Health education and face-to-face inquiry.

Participants will be recruited by trained and experienced fieldworkers from six cities: Shenzhen, Beijing, hangzhou, Zhengzhou, Qingdao and Hangzhou. Recruitment methods include: (1) recruiting eligible MSM individuals presenting at the participating hospitals; (2) recruiting MSM in the community through community-based organizations (CBOs); (3) peer referral.

#### Ethics approval

All participants must provide informed consent before enrollment. Rresearch assistants will fully explain the study’s purpose, procedures, potential risks, and benefits, emphasizing its voluntary nature and confirming that refusal will have no consequences. Recruitment is finalized only after consent is signed. To ensure confidentiality, each participant’s data will be strictly protected. Meanwhile, all research staff must complete training and sign confidentiality agreements prior to project commencement. Questionnaires will maintain anonymity by using unique identification codes, which will be matched during follow-up, with the code list securely managed by designated personnel. Data collected via the online platform Jinshuju (https://jinshuju.net/home) will exclude personally identifiable information. Only principal investigators can view or export the data. To prevent data loss, a full backup will be stored on the principal investigator’s hard drive monthly.

Participants must review and sign consent forms, permitting collection of location and IP data. After submission, they will receive a non-editable report to verify answers. One revision is allowed upon auditor approval, with timestamps recording changes to ensure accuracy.

For each completed follow-up visit, participants receive a 50 yuan compensation for lost work time, with a maximum compensation of 250 yuan.

The protocol has been approved by the Peking University ShenZhen Hopital Ethics Committee(PUSHEC) and registered at *Chinese Clinical Trial Registry* (ChiCTR2500111280). Any subsequent amendments will be submitted to PUSHEC for review. Once approved, corresponding modifications will be implemented on *Chinese Clinical Trial Registry*.

### Sample Size

This study is a three arms, multicenter, randomized, open-lable RCT. Primary outcome is difference of good PrEP adherence in PrEP-bot-SPB and HEF interventions. Previous study showed that the percentage of PrEP adherence in HEF interventions was 70.9%. We assume that 70% of participants in the control group maintain good PrEP adherence (PrEP adherence ≥90%) at month 3 of the project. The difference between the intervention and control groups is 20%, With the consideration of 90% power, 1-sided significance level of .05. Using PASS 15.0, a sample size of 80 per group was calculated. Accounting for a 20% dropout rate, at least 100 participants per group (total N ≥ 300) are required.

#### The Baseline Survey and Random Allocation Process

##### Randomization

After baseline survey, participants will be equally randomized into the intervention or control groups. Allocation concealment is implemented during the randomization process, ensuring that neither the investigators nor the participants can foresee the group assignment. The specific procedure is as follows: A randomization sequence will be generated using the randomization module in SPSS 26.0. A research staff member without involvement in recruitment or baseline survey will seal this sequence within sequentially numbered, opaque, sealed envelopes. Each eligible participant will be assigned a unique identification number. The envelope corresponding to each participant’s number will then be distributed to them, thereby implementing allocation concealment.

##### Blind Method

As participants will aware of their assigned intervention after randomization, this study employs an open-label design. However, the research staffs in charge of grouping has no idea about the specific grouping situation.

### The Control Groups

#### HEF

After randomization, trained researchers will provide a health education pamphlet introducing PrEP and knowledge related to PrEP medication adherence at baseline. During follow-up visit at months 3, 6, 9, and 12, trained researchers will inquire face-to-face about participants’ PrEP adherence and PrEP-related health check-up behaviors over the past three months. For participants reporting missed doses or incomplete check-ups, researchers will explain the necessity and potential hazards of skipped check-ups, and provide available check-up locations.

### The Intervention Groups

This study comprises two intervention groups: the PrEP-bot group and the PrEP-bot-SPB group. Participants in the PrEP-bot group will be granted access to a PrEP-bot agent through WeChat. The agent will: 1) generate personalized PrEP medication plans; 2) provide medication reminders and PrEP-related health check-ups notifications; 3) inquire reasons about missed doses to deliver tailored interventions; 4) answer participant questions about PrEP using guideline-based knowledge. Participants in the PrEP-bot-SPB group will not only receive a PrEP-bot agent through WeChat but also receive a SPB that delivers PrEP medication reminders via vibration and auditory alerts. PrEP-bot agent will obtain PrEP medication plans and medication adherence information from SPB through an application programming interface (API). Participants will receive both the SPB intervention and the PrEP-bot intervention.

#### Intervention Procedure

##### Install Intervention System

After randomization, participants in the intervention groups will add the PrEP-bot agent using WeChat with the help of trained researchers. The agent can identify the identities of participants through WeChat IDs. This process does not involve the display and interaction of participants’ personal identity information, ensuring the confidentiality of their details. The PrEP-bot and SPB systems will be interconnected via API to enable medication adherence data exchange.

Furthermore, trained researchers will distribute SPBs to participants in the PrEP-bot-SPB group; then demonstrate SPB operation procedures, safety precautions, and configuration settings to participants; supply instructional videos for self-directed SPB adjustment; and offer ongoing technical support during device usage. After receiving the SPB, participants scan the device’s barcode via WeChat and follow the instructions to link their phone number to the Apei Health WeChat service account, link the SPB to the corresponding WeChat mini-program. This completes the registration process and establishes a unique pairing between each participant and their SPB. The entire process is anonymous, guaranteeing the confidentiality of participants’ personal details. Participants may contact the researchers at any time for support and assistance with the SPB.

#### The PrEP-bot Intervention Procedure

##### The PrEP-bot Intervention Procedure for PrEP Adherence

The intervention process is accomplished through prompt engineering and agent technology, enabling PrEP-bot to make autonomous decisions in a pre-defined workflow. It can independently determine the medication stage of the users and provide reminders and medication guidance. After participants add PrEP-bot, it will ask the user about the high-risk sexual behavior plan and generate guideline-based PrEP regimens (daily or event-driven PrEP), which will always be present in the memory module and context prompts of PrEP-bot. The PrEP-bot will deliver scheduled PrEP reminders at scheduled times. In this study, a missed dose is defined as not taking PrEP 2 hours after the scheduled time. According to the instructions for HIV medication, a missed dose can be taken up to 18 hours after the scheduled time. If participant who miss a dose, he could take it within 18 hours. Otherwise, wait until the next scheduled dose. When the user is found to have missed dose, the PrEP-bot will initiates follow-up inquiries at, 6-, 12-, and 18-hour intervals post-scheduled time. Reminders cease immediately upon users confirmation of dose intake, proceeding to the next scheduled dose. For unresponsive users, the PrEP-bot will deliver educational content regarding non-adherence consequences, solicits reasons for missing the dose, and provides personalized intervention messages. All reminders for the current dose automatically terminate 18 hours post-scheduled time, advancing to subsequent doses. The researchers managing the PrEP-bot will track missed doses and activate a four-tier response mechanism based on the number of missed doses of the users. Specifically, after six missed doses, the PrEP-bot will send WeChat articles or videos containing PrEP medication guidelines and expert consensus. For up to eight missed doses, family members of the user will be notified to assist with medication reminders. If eight or more doses are missed, the trained researchers will counsel the user personally to resume taking the medication.

##### PrEP-bot Intervention Procedure for PrEP-related health check-ups

The PrEP-bot will ask users for PrEP-related health check-ups information and generates personalized testing schedules. Users will receive health check-ups reminders during the 7-day pre- and post-appointment windows, with confirmation prompts upon completion. Reminder cycles conclude upon user response and reset for subsequent health check-ups. Otherwise, initiating the four-tier response mechanism.

##### PrEP-bot Answer Users question about PrEP

The user can ask the PrEP-bot questions. The system of PrEP-bot processes user inquiries through keyword recognition, delivering preconfigured responses containing matched terms to enable precision interventions. PrEP-bot can independently distinguish between standardized consultations and complex medical issues, and transfer complex medical issues to the trained researchers.

#### The PrEP-bot-SPB Intervention Procedure

##### SPB Intervention Procedure for PrEP Adherence

SPB reminders PrEP participants through two ways. ①Self-reminder model: users can set a PrEP reminder plan on Aipei health, a wechat mini-program of SPB. After setting the plan, both SPB and Aipei health service (a wechat service account) issue alerts at scheduled time. Opening the SPB within 2 hours pre-dose to 18 hours post-dose registers as PrEP taken. If the SPB remains unopened, it will ring again after 30 minutes and 60 minutes respectively. ②System background monitoring and remind mode: Chip on the SPB automatically transmits real-time adherence data to cloud servers, trained administrators will identify missed doses and send reminder message to users. Moreover, participants will receive placement guidance (e.g., “maintain SPB on dining tables”, “carry the SPB or other convenient medicine containers when you go out”) to minimize forgetfulness-related omissions.

Participants in PrEP-bot-SPB group will receive both the SPB and PrEP-bot agent interventions. The SPB is connected to the PrEP-bot via API to deliver users’ PrEP adherence data. Transmitted parameters include: SPB lid status (open/closed), PrEP reminder plan. Once the PrEP-bot get users PrEP reminder plan, it will reminder users at the scheduled time. Opening the SPB within a ±2 hour window of scheduled dosing registers as on-time PrEP intake. This adherence signal triggers immediate cessation of corresponding dose reminders of PrEP-bot through the API. Otherwise, the PrEP-bot will assumes the dose as missed and subsequently send a medication reminder at 2, 6, 12,18 hours after the scheduled time, until either the pill box is opened or 18 hours have passed since the scheduled dose time.

##### Intervention Termination

Participants will be withdrawn from the intervention under the following circumstances: 1) completion of the follow-up period; 2) testing positive on an HIV self-test during follow-up and subsequently being confirmed HIV-positive upon confirmatory testing; 3) experiencing severe adverse events and being deemed unsuitable to continue participation by professional researchers or physicians; 4) voluntary withdrawal from the study for personal reasons.

##### Intervention Adjustment

Based on the PrEP expert consensus, participants who frequently miss doses under the event-driven dosing regimen will be advised to switch to the daily dosing regimen.

### Interventions

#### Overview of the SPB

The SPB interfaces with two companion platforms, the Aipei Health WeChat Mini Program and Service Account. The mini-program enables configuration or resetting of medication schedules, adjustment of SPB alert parameters (volume levels and vibration toggles), and monitoring of battery status. An embedded sensor chip within the SPB continuously monitors operational status. The Service Account primarily delivers medication reminders while enabling users to review dosing schedules and adherence records.

#### Function of the SPB

All configuration commands issued via the mini-program are detected by the sensor chip and subsequently transmitted to cloud servers. After receiving the SPB, users can establish medication schedules, adjust volume settings, and configure vibration modes via the mini-program. The SPB triggers audiovisual alerts while the Service Account simultaneously pushes medication reminders at scheduled dosing times. Lid activation during administration windows is registered as a confirmed dosing event. When miss dose is detected, supplemental alerts are initiated by the SPB 30 minutes post-scheduled administration. Upon regimen completion, reminder functionalities are deactivated through mini-program reset protocols, with subsequent reactivation occurring when new PrEP dosing requirements emerge.

### Development of the PrEP-bot

#### Overview

The research teams from *Tsinghua University and Chinese Academy of Sciences* and *University of Chinese Academy of Sciences* develop the chatbots. The chatbots integrates with WeChat through its public Web API services. Based on the user’s high-risk sexual behavior plans and type of PrEP use (daily or event-driven PrEP), the PrEP-bot will generate a corresponding medication plan. They can formulate a PrEP-related health check-ups plan according to the user’s pre-enrollment health check-ups history. The PrEP-bots will send medication reminders and health check-ups notifications at scheduled times. Additionally, it will be capable of answering user questions related to PrEP.

#### PrEP-bot Development Process

The PrEP-bots are developed based on health behavior theories. Our team refers to PrEP consensus documents and guidelines and collects PrEP-related materials from community-based organizations (CBOs). Specific medication reminder processes are established for both daily PrEP and event-driven PrEP, along with a dedicated health check-ups reminder process for PrEP-related health check-ups. Reminder rules are configured within agents. To provide personalized interventions for missed doses, relevant information on missed and supplemental doses is compiled into a question-answer pairs (Q&A) knowledge base. When a user misses a dose, chatbots will proactively inquire about the reason. After receiving the user’s response, PrEP-bots will identify keywords and send corresponding content. Beyond medication reminders and missed-dose interventions, other PrEP-related knowledge is incorporated into the Q&A knowledge base, enabling PrEP-bots to respond to inquiries about PrEP and HIV.

All Q&A in the knowledge base will be integrated with agents system using knowledge retrieval technology. This will supplement PrEP-bots with knowledge and solutions for common concerns and misconceptions, enabling they to provide reliable responses through Retrieval-Augmented Generation (RAG). Meanwhile,The research team will weekly refine the PrEP-bot based on issues identified during user interactions to improve its accuracy.

#### Functionality of the PrEP-bot

The functionalities of PrEP-bots primarily include: 1) Intent recognition: accurately identifying user requests, such as adjusting medication or health check-up schedules, or answering health-related inquiries; 2) User memory: recording key user information, medication history, and recent conversation logs to enable personalized services; 3) Medication reminders: developing a PrEP medication schedule for users and sending reminders at preset times; 4) Personalized missed-dose intervention: initiating a dialogue when a missed dose is detected, inquiring about the reason, and delivering tailored intervention messages based on the specific cause; 5) Health check-up reminders: creating a PrEP-related health screening plan and sending reminders at scheduled times; 6) Health consultation: engaging in dialogue with users to answer their questions; 7) Emotional regulation: expressing positive reinforcement for consistent PrEP adherence and showing empathy toward users’ concerns about medication. Shown in figure 2A, figure 2B.

**Figure 2A.**
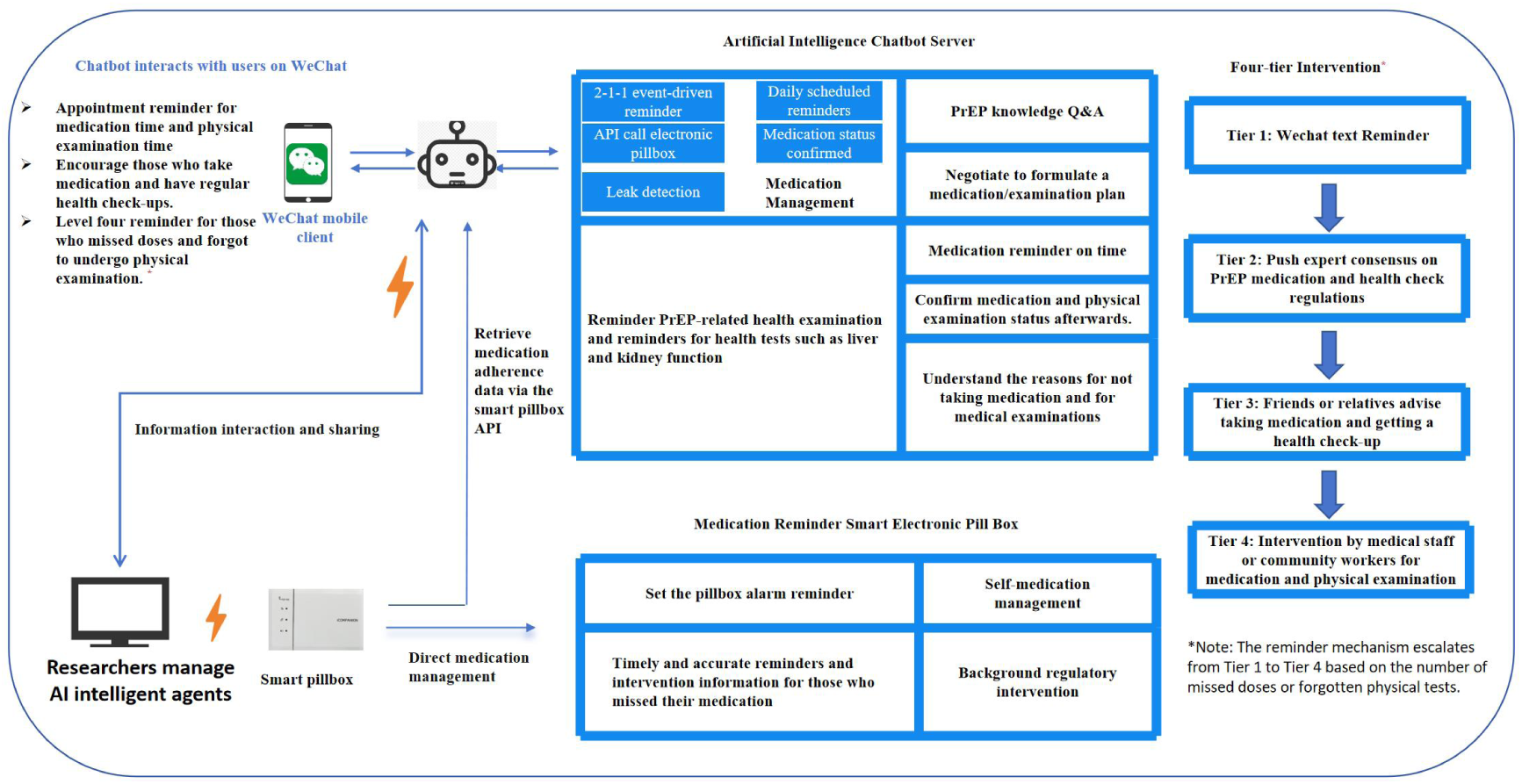
Intervention diagram for MSM’s HIV PrEP medication adherence and physical test behavior through AI-Chatbox + smartpillbox. MSM: men who have sex with men; PrEP: Pre-exposure prophylaxis

**Figure 2B.**
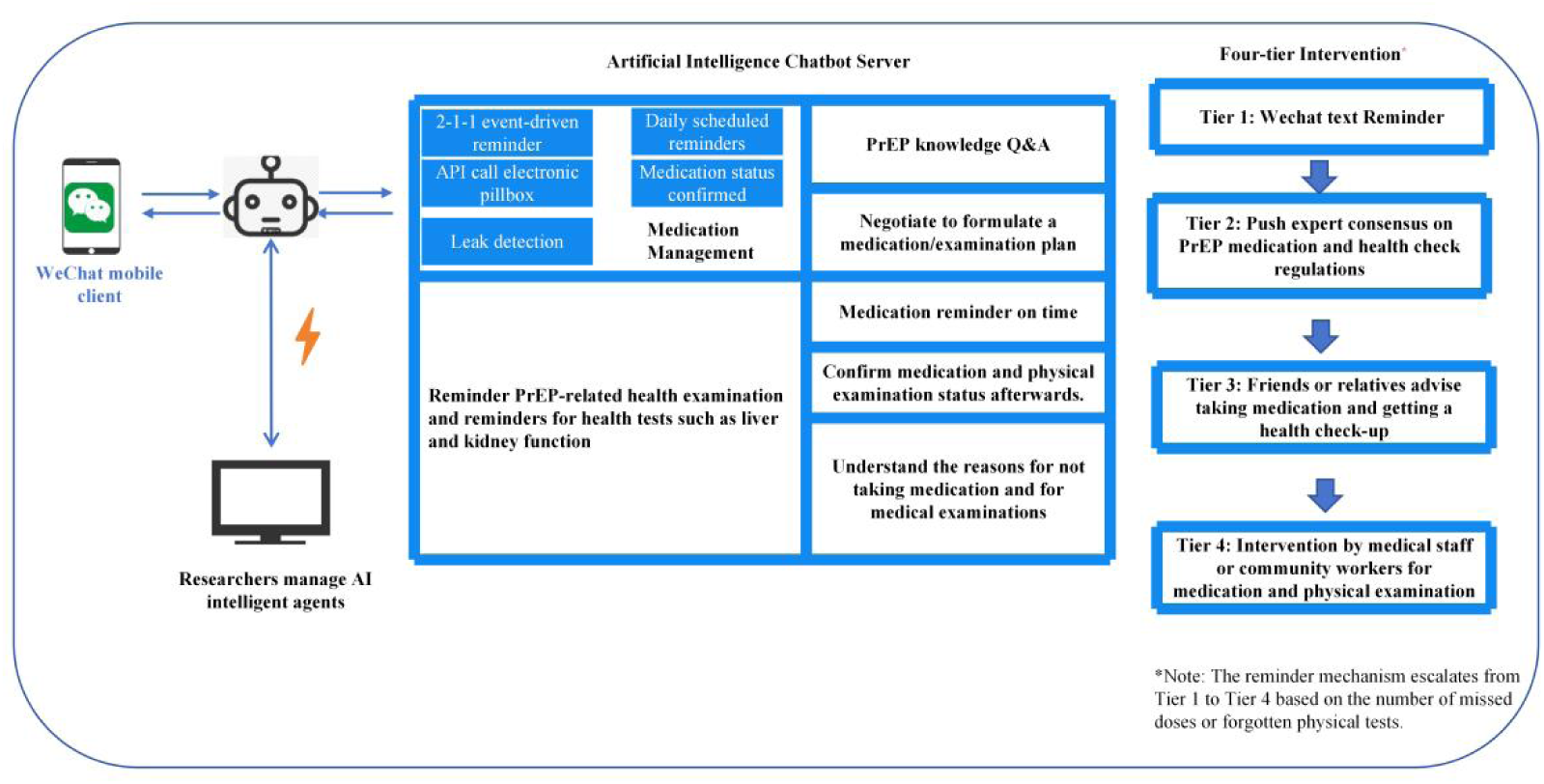
Intervention diagram for MSM’s HIV PrEP medication adherence and physical test behavior through AI-Chatbox. MSM: men who have sex with men; PrEP: Pre-exposure prophylaxis

When user inquiries exceed the knowledge base, PrEP-bots will trigger an alert and refer the user to a trained research staff member for further assistance. When users send messages unrelated to PrEP, chatbots will reply: “I am unable to answer this type of question,” followed by the prompt: “If you have any questions about PrEP, HIV prevention, or health-related issues, feel free to consult me anytime.”

#### Measurements

##### Primary outcome

The primary outcome is PrEP adherence at 3 month of follow-up, which will be measured through three different methods: (1) Self-reported; (2) MMAS-8; (3) TFV blood concentrations. Outcomes will be measured by trained administrators.

**Self-reported PrEP adherence** will be calculated by dividing the difference between the number of pills that should have been taken and the number of pills missed by the number of pills that should have been taken. The average number of pills missed per week over the past three months will be recalled by each participant through a self-completed online questionnaire. 90% PrEP adherence is classified as “good” PrEP adherence.

**PrEP adherence based on MMAS-8** will be measured by MMAS-8. MMAS-8 consists 8-items. For items 1-7, response options are “yes” or “no,” with a score of 1 assigned for “no” and 0 for “yes.” Item 8 have five response options: “never,” “rarely,” “sometimes,” “often,” and “all the time,” scored using a 5-point Likert scale as 1.00, 0.75, 0.50, 0.25, and 0.00, respectively. The total MMAS-8 score ranged from 0 to 8, with scores <6 defined as low adherence, 6-8 as medium adherence, and 8 as high adherence.

**PrEP adherence based on TFV blood concentration** will be evaluated on blood samples from DBS and 3ml venous blood, with 5% of participants in each group being measured. TFV blood concentration ≥35.5 ng/mL are considered indicative of ≥90% PrEP adherence and classified as “good” PrEP adherence.

###### Secondary Outcomes

Secondary outcomes include: 1) PrEP adherence at the 6-th, 9-th and 12-th month of follow-up; 2) HIV incidence rate; 3) completion rate of PrEP-related health check-ups; 4) effectiveness, feasibility, acceptability, and user satisfaction of the PrEP-bot; 5) the consistency between the proportion of participants with good PrEP adherence (defined as ≥90%) based on self-report and the MMAS-8, and the proportion with good adherence (defined as TFV ≥35.5 ng/mL) based on DBS; 6) the interventions effects of the PrEP-bot-SPB and the PrEP-bot on daily PrEP and event-driven PrEP, respectively; 7) the health economics effect of the PrEP-bot-SPB intervention compared with the traditional PrEP intervention approach.

HIV incidence rate is calculated as the number of newly detected and confirmed HIV cases divided by the number of participants who were HIV-negative at baseline within the cohort at each follow-up interval. participant’s fingertip blood sample will be collected to determine the HIV infection status using an HIV self-test kit. If the self-test kit indicates a suspected HIV positive result, the participant will be referred to a local hospital or CDC for confirmation of the HIV infection status through enzyme-linked immunosorbent assay and Western blot. Participants confirmed to be HIV positive will be withdrawn from the intervention. Additionally, during the follow-up period, if necessary, researchers can mail HIV self-test kits to participants at any time.

The effectiveness of PrEP-bots will be assessed by comparing PrEP adherence between participants in PrEP-bots group and those in the non-PrEP-bot group. A greater difference indicates higher effectiveness of PrEP-bots.

Feasibility will be evaluated using a feasibility assessment scale of chatbots. The scale comprised four domains: overall quality of PrEP-bots, satisfaction scale, intent to continue using PrEP-bots, willingness to refer PrEP-bots to others. Each domain was scored on a 1-10 scale, with higher scores indicating better feasibility.

Acceptability will be measured using the standardized System Usability Scale (SUS) and an Adjusted Chatbot Usability Scale (ACUS). Each scale consisted of 10 items, scored on a 1-5 point scale. Higher scores on these scales indicated better acceptability. Scores on SUS are also used to reflect user satisfaction, with higher scores indicating greater satisfaction.

PrEP-related health check-ups rate will be defined as the proportion of participants who completed scheduled PrEP-related health check-ups among all participants at each group. A questionnaire will be filled out to ensure whether the participants have health check-ups for HIV antibodies, liver and kidney functions, HBV infection indicators and STD tests (syphilis, gonorrhea and chlamydia infection) on time.

###### Baseline survey

The content of the baseline survey questionnaire include: sociodemographic information: age, educational level, marital status, personal monthly income, phone number of the participant, phone number of a family member for reminder purposes (PrEP intake and health check-ups); sexual behavior characteristics information: ways of finding sexual partners, types of sexual partners, sexual roles, number of sexual partners in the past six months, condom use, number of unprotected anal sex with casual partners; Types of PrEP; HIV risk perception, HIV awareness rate and HIV counseling rate; PrEP-related knowledge and attitudes, PrEP missed doses and factors affecting medication adherence, etc.

###### Adverse Events

The adverse events of interest in this study are potential side effects associated with PrEP use. Short-term side effects may include nausea, headache, diarrhea, and fatigue, while long-term risks may involve mild renal function decline, lactic acidosis, decreased bone mineral density, weight loss, and electrolyte imbalances. Although these adverse events are not caused by the study interventions, they will be actively monitored by our team. Participants will be advised to seek medical care or temporarily discontinue PrEP if needed to ensure safety.

###### Safety Outcomes

Safety outcomes include the observation and documentation of adverse events. During the trial, any participant experiencing a severe adverse event will have the event recorded in detail, including the time of onset, severity, duration, actions taken, and outcome. Participants withdrawing due to adverse events will be followed until resolution. The number of discontinuations due to severe adverse events will be reported after follow-up.

##### Follow-up

###### Overview

Participants will be followed for 12 months, with combined online and face-to-face assessments at 3, 6, 9, and 12 months. The outcomes are distinguished between short-term and medium-to-long-term effects.

###### Offline follow-up

Face-to-face follow-up visits will be conducted for each of the three study groups every three 3 months. During each visit, questionnaires will be administered to collect participants’ PrEP adherence data based on self-reporting and the MMAS-8, the completion rate of PrEP-related health check-ups; the data of high-risk sexual behavior (number of sexual partners, frequency of sexual activity, condom use rate). Participants will also be asked to evaluate the effectiveness, feasibility, acceptability, and satisfaction of using the PrEP-bot. Peripheral blood samples will be collected from participants for HIV antibody testing. For those who were selected for TFV blood concentration, 3ml venous blood samples will be collected from participants to determine PrEP adherence based on serum. For the traditional PrEP intervention group, researchers also asked participants about the reasons for non-adherence to PrEP and PrEP-related health check-ups, informed them of the importance and necessity of taking medication on time and undergoing health check-ups, and provided them with the locations and times for health check-ups.

###### Online follow-up

To prevent users from being lost to follow-up, an online wechat/phone follow-up will be conducted for the three groups between two offline follow-ups to understand the participants self-reported PrEP adherence and PrEP-related health check-ups behaviors.

###### Process Evaluation

At month 3, short-term effects including PrEP adherence, HIV incidence, PrEP-related health check-ups completion rates and the effectiveness, feasibility, acceptability, and user satisfaction of PrEP-bots will be analyzed. Medium-to-long-term effects will be assessed at the 6-month, 9-month and 12-month follow-ups.

##### Statistical analysis

Particpants who attend the follw-up will be analysis, with 5% of them will be analysis for PrEP adherence based on DBS and venous blood. All analyses followed the intention-to-treat (ITT) and per protocol set (PPS) analysis. Multiple imputation was used to handle missing data. Chi-square tests and t-tests will be employed to compare baseline characteristics among the three groups. Relative risk reduction, absolute risk reduction, and their 95% confidence intervals will be used to compare differences in PrEP adherence (PrEP adherence ≥90% base on self repoted, MMAS-8, DBS and venous blood), HIV infection rate, and PrEP-related health check-ups completion rates between the three groups. Means and standard deviations will be used to describe the effectiveness, feasibility, acceptability, and user satisfaction of the PrEP-bot. A mixed-effects model will be applied to analyze differences in PrEP adherence over time among the four groups. ROC curves and Kappa coefficients will be utilized to evaluate the agreement between the proportion of participants with good medication adherence (≥90%) as measured by self-report and the MMAS-8, and the proportion with good PrEP adherence (TFV ≥35.5 ng/mL) as determined by DBS and venous blood. Subgroup analysis will be conducted to compare the difference in intervention effects between daily PrEP and event-driven PrEP. Data will be analyzed using SPSS 27.0, statistical significance is set at the *P*<.05 level (2-sided).

## Clinically and politically significant

The efficacy of PrEP in preventing HIV has been robustly demonstrated, underscoring the necessity of its widespread implementation among MSM. However, PrEP’s preventive efficacy is contingent upon adherence, making consistent and correct usage a critical determinant of HIV prevention. Although multiple interventions to enhance PrEP adherence exist, their impact remains suboptimal due to limitations such as delayed feedback, lack of personalization, and substantial resource demands. Consequently, significant room for improvement persists in PrEP adherence among MSM populations. In addition, the regular PrEP-related health check-ups rates of the population taking PrEP is also a problem that needs attention.

To our knowledge, this represents the first attempt to employ an AI-chatbot to deliver medication reminders, health monitoring alerts, and informational support for MSM using PrEP. This RCT will compare the efficacy of PrEP-bot-SPB against a traditional PrEP adherence strategy—HEF—in increasing PrEP adherence, enhancing HIV prevention effectiveness and promoting PrEP-related health check-ups among MSM. Should the PrEP-bot-SPB prove potential non-inferiority than HEF in improving PrEP adherence, regular PrEP-related health check-ups and HIV prevention outcomes, it may address existing gaps in PrEP adherence and regular health check-upsinterventions for MSM, meanwhile, save a great deal of manpower and material resources.

Leveraging real-time delivery, precision personalization, and the scalability of AI technology, The PrEP-bot-SPB intervention holds promise for adoption by governmental organizations and CBOs in China. This intervention has the potential to overcome barriers inherent in traditional interventions, thereby significantly improving PrEP adherence and regular PrEP-related health check-ups among MSM and reducing HIV incidence in this population.

While this trial will yield essential insights for policy formulation, several limitations warrant acknowledgment. First, participants will be recruited through convenience sampling, which may introduce selection bias. Second, Cautions should be taken when generalizing findings to the broader MSM population in China. Third, collecting characteristics of MSM who decline to participation is challenging, further potentiating selection bias.

## Conclusion

This study will generate critical research and policy implications regarding PrEP-bot applications in PrEP adherence and health check-ups interventions. If the PrEP-bot demonstrates not inferior than HEF, its real-time precision, low resource requirements, and automated nature could facilitate seamless integration into existing PrEP adherence frameworks. The intervention holds the potential to overcome traditional barriers, thereby enhancing PrEP adherence and PrEP-related health check-ups among MSM and improving control of HIV transmission in this key population.

## Supporting information

The SPIRIT recommendations detail is shown in supplementary file

## List of Abbreviations

ACUS: Adjusted Chatbot Usability Scale
AI: Artificial Intelligence
API: Application Programming Interface
CBOs: Community-Based Organizations
DBS: Dried Blood Spot
Gen-AI: Generative Artificial Intelligence
HBV: Hepatitis B Virus
HE: Health education
HEF: Health education and face-to-face inquiry
HIV: Human Immunodeficiency Virus
ITT: Intention-to-Treat
LLM: Large Language Model
MAR: Medication Reminder Assistant
MMAS-8: 8-item Morisky Medication Adherence Scale
MSM: Men Who Have Sex with Men
PPS: Per Protocol Set
PrEP: Pre-exposure Prophylaxis
PrEP-bot: PrEP Chatbot
PrEP-bot-SPB: PrEP-bot + Smart Pillbox
PUSHEC: Peking University ShenZhen Hopital Ethics Committee
RAG: Retrieval-Augmented Generation
RCT: Randomized Controlled Trial
ROC: Receiver Operating Characteristic
SPB: Smart Pillbox
STI: Sexually Transmitted Infection
SUS: System Usability Scale
TFV: Tenofovir
UNAIDS: Joint United Nations Programme on HIV/AIDS
WHO: World Health Organization

## Acknowledgements

The authors appreciate the help of CBOs in recruiment of participants, we also thank the research teams at Tsinghua University and Chinese Academy of Sciences and for their effoets in the development of interventions.

## Article Information

### Contributions

RBZ, ZLZ, HHY, and JJX designed this protocol, initiated the research plan and designed the measurements. RBZ, ZLZ, HHY, YXW and HL prepared the materials for the development of AI-chatbots. JJX and XJH acquired the funding for this study. RBZ edited the manuscript. ZLZ, HHY, YXW, HL, YJJ, JJX, GYW, NZ, HFH, XJH, XLZ and QRS critically reviewed the manuscript, providing insights on important intellectual content, and approved the final version. All the authors reviewed and agreed the final manuscript.

## Availability of data and materials

Data sharing is not applicable to this article as no datasets were generated or analysed during the current study.

## Ethics and approval and consent to participate

Written informed consent will be provided to all study participants before take part in this study. The study was approved by the Peking University Shenzhen Hospital {Peking University Shenzhen Hospital Ethics Review (Research) [2025] No. (230-Rev.3)}.

## Consent for publication

Not applicable.

